# COVID-19 Outcomes in 4712 consecutively confirmed SARS-CoV2 cases in the city of Madrid

**DOI:** 10.1101/2020.05.22.20109850

**Authors:** Sarah Heili-Frades, Pablo Minguez, Ignacio Mahillo Fernández, Tomás Prieto-Rumeau, Antonio Herrero González, Lorena de la Fuente, María Jesús Rodríguez Nieto, Germán Peces-Barba Romero, Mario Peces-Barba, María del Pilar Carballosa de Miguel, Itziar Fernández Ormaechea, Alba Naya prieto, Farah Ezzine de Blas, Luis Jiménez Hiscock, Cesar Perez Calvo, Arnoldo Santos, Luis Enrique Muñoz Alameda, Fredeswinda Romero Bueno, Miguel Górgolas Hernández-Mora, Alfonso Cabello Úbeda, Beatriz Álvarez Álvarez, Elizabet Petkova, Nerea Carrasco, Dolores Martín Ríos, Nicolás González Mangado, Olga Sánchez Pernaute, and the COVID FJD-TEAM

## Abstract

There is limited information describing features and outcomes of patients requiring hospitalization for COVID19 disease and still no treatments have clearly demonstrated efficacy. Demographics and clinical variables on admission, as well as laboratory markers and therapeutic interventions were extracted from electronic Clinical Records (eCR) in 4712 SARS-CoV2 infected patients attending 4 public Hospitals in Madrid. Patients were stratified according to age and stage of severity. Using multivariate logistic regression analysis, cut-off points that best discriminated mortality were obtained for each of the studied variables. Principal components analysis and a neural network (NN) algorithm were applied.

A high mortality incidence associated to age >70, comorbidities (hypertension, neurological disorders and diabetes), altered vitals such as fever, heart rhythm disturbances or elevated systolic blood pressure, and alterations in several laboratory tests. Remarkably, analysis of therapeutic options either taken individually or in combination drew a universal relationship between the use of Cyclosporine A and better outcomes as also a benefit of tocilizumab and/or corticosteroids in critically ill patients.

We present a large Spanish population-based study addressing factors influencing survival in current SARS CoV2 pandemic, with particular emphasis on the effectivity of treatments. In addition, we have generated an NN capable of identifying severity predictors of SARS CoV2. A rapid extraction and management of data protocol from eCR and artificial intelligence in-house implementations allowed us to perform almost real time monitoring of the outbreak evolution.

## INTRODUCTION

The new coronavirus pandemic was confirmed to have spread to Spain on 31 st January 2020, when a German tourist tested positive for SARS-CoV-2 in La Gomera, Canary Islands. Post-hoc genetic analysis showed that at least 15 strains of the virus had been imported and community transmission begun by mid-February^1^. By 13th March, cases had been confirmed in all of the state 50 provinces.

A state of alarm and national lockdown was imposed on 14th March. On 29th March it was announced that, beginning the following day, all non-essential workers were to stay at home for the following 14 days. By late March, Madrid had recorded the highest number of cases and deaths in the country. On 25th March 2020, the death toll in Spain surpassed the one reported in Mainland China, while only deaths in Italy remained higher. As of 9th May 2020, there have been 223,578 confirmed cases with 133,952 recoveries and 26,478 deaths in Spain. Regarding the course and outcomes of SARS-CoV-2 pneumonia, there are already population-based data from China ^2^, Singapore^3^, Italy^4^ and recently the United States of America^5^. This work aims to present a comprehensive analysis of COVID-19 risk factors including vital signs, comorbidities, and biomarkers, as well as treatment effectiveness using machine-learning and classifying mortality predictors of SARS CoV-2 based on a large population cohort from 4 public Hospitals in Madrid.

## MATERIAL AND METHODS

### Study Population

A total of 4712 patients attending any of the 4 centers participating in the study, from January 31st 2020 to April 17th 2020 were included. All patients had confirmed SARS-CoV-2 infection by a positive result on polymerase chain reaction testing. Participating hospitals shared a common clinical practice guidance for admission, follow-up and inpatient management of SARS-CoV2 infection (**detailed in Supplementary Table 1**). De-identified medical records were confidentially collected from Microsoft Sql Integration Services (SSIS) reporting database and analysed using R version 3.5.2 (R Project for Statistical Computing; R Foundation), MATLAB 9.7-R2019b and IBM SPSS Statistics-25. Approval from the Institutional Ethics Committee was obtained.

### Data Collection and baseline variables included in the study

Demographics, baseline comorbidities, vital signs, biomarkers, use of treatments and outcomes were extracted from clinical charts. Tobacco consumption was analysed and multivariate logistic regression models were performed testing potential interactions between tobacco and comorbidities.

### Patient stratification, biomarkers measure selection and outcome codification

All data were adjusted for age, taken as a continuous variable. In addition, patients were stratified into 5 age subgroups (0-16, 17-30, 31-50, 51-70 and 71-100) and in the range 31-100 separately. Biomarkers with > 50% of non-missing data at first day of hospitalization were included in the analysis. The effect of treatments was studied in those patients receiving at least one drug from the clinical protocol (N=2776). Untreated patients were mild cases discharged from the emergency department. Patients were classified according to severity of the condition at the time of data collection (17/04/2020), specifically recording fatal outcomes and ICU admission. Thus, analysis and predictions were made using the following classification frameworks: 1) deceased patients and those admitted to ICU versus rest, 2) deceased versus rest, and 3) deceased versus ICU admitted.

### Missing Data Imputation

Parameters with a high percentage (>50%) of missing values were excluded from the analysis. Missing data for biomarkers and vital signs were imputed using average value within age ranges considered in each analysis.

### Neural Network And Data Re-Sampling

The neural network (NN) model was built using the caret R package (https://CRAN.R-project.org/package=caret). We used “nnet” method, and implemented an automatic selection of: i) the optimal number of units per hidden layer (1 to 5), and ii) the optimal value for the regularization parameter to avoid over-fitting (0.1 to 0.5 in increments of 0.1 units). We used “twoClassSummary” method to compute sensitivity, specificity and the area under the ROC curve. This NN algorithm was performed 10 times over a matrix of re-sampled patients. Re-sampled matrices were composed of patients with severe status (outcome = 1) and a random selection of the same number of patients with less severe status (outcome = 0). For each re-sampled matrix, we performed a 10-fold cross validation where randomly used 90% of patients as the training dataset and the remaining (10%) as the testing dataset. A total of 100 NN models were performed and a ROC curve is calculated for the set of predictions and real values using the 10-fold cross validation. Our final AUC, accuracy, sensitivity and specificity were calculated as the mean of the 100 NN models performed in total. Our R script is available free at: https://github.com/pminguez/MachineLearning4UnbalancedData/blob/master/nn4covid19.R.

### Principal Component Analysis

Principal component analysis (PCA) was performed on the matrices with missing data imputation, centered and scaled, using the prcomp and factoextra R libraries. Elipses represent 95% of the data. PCA analyses were performed over the same scenarios chosen to perform the NN (see above section).

### Fisher exact test analysis

For binary type of variables, which included comorbidities, treatments and gender, odds ratios for each of the two severity classes in each comparison were calculated, contingency 2×2 tables were obtained and significance was calculated with 2-tailed Fisher test. P values were adjusted using FDR and adjusted p values <0.05 were taken as significant. Graphics were built sorting variables by relative odds ratios (1/odds ratio when <1).

### Wilcoxon test analysis

For non-binary variables, comprising vital signs and biomarkers, a non-parametric Wilcoxon rank sum test was performed over the set of values of each variable for the two severity states. P-values were adjusted by FDR and FDR<0.05 were considered as significant. Means of the two sets of values for each variable was calculated to assign a direction of the test.

### Classification and Cut-Off Values Based on vital signs

Logistic Regression models were used to obtain the vital signs cut-off values and to discriminate those, which were related to an unfavorable evolution. Using ROC curves and Youden’s J statistic we obtained the cut-off values. We defined a binary classification for each parameter based on the cut-off points and selected parameters (p-value<0.2) to feed a multivariate logistic regression model. ROC curves for the model were calculated on 10-fold cross validation, and AUC, accuracy, sensitivity and specificity were averaged across the 10 ROC curves. The goodness of fit of the model was evaluated by the Hosmer and Lemeshow test.

### Analysis of the effect of drugs on survival in this cohort of patients

Two models were used for this approach.

**Model 1: First**, a logistic regression was performed to individually determine the relationship of each of the treatments included in the local interim clinical guidance protocol with the survival of the patients. We considered 26-predictor variables *X_i_* for each of the 26 treatments. A value of 1 was assigned for the variable *X_i_* if the corresponding treatment had been applied, whereas a value of 0 represented non-exposure. The Multinomial Logistic Regression model had the form

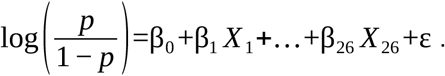

Hence, the approximation for *p* was

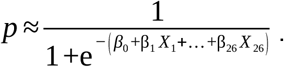

We use the Multinomial Logistic Regression fitting mrnfit function in Matlab for the sample of 4712 individuals.

**Model 2: Second**, we applied our NN based algorythm to detect treatments predicting success and failure both in the global population of 4712 (which included a 40% of mild cases not receiving specific treatment, (table 1, suppl table 2) and in the 2739 patients (60% of global cohort, table 1) who were admitted to hospital. In the latter cohort, the effect of the medications on the different age subgroups was determined.

**Table 1.**
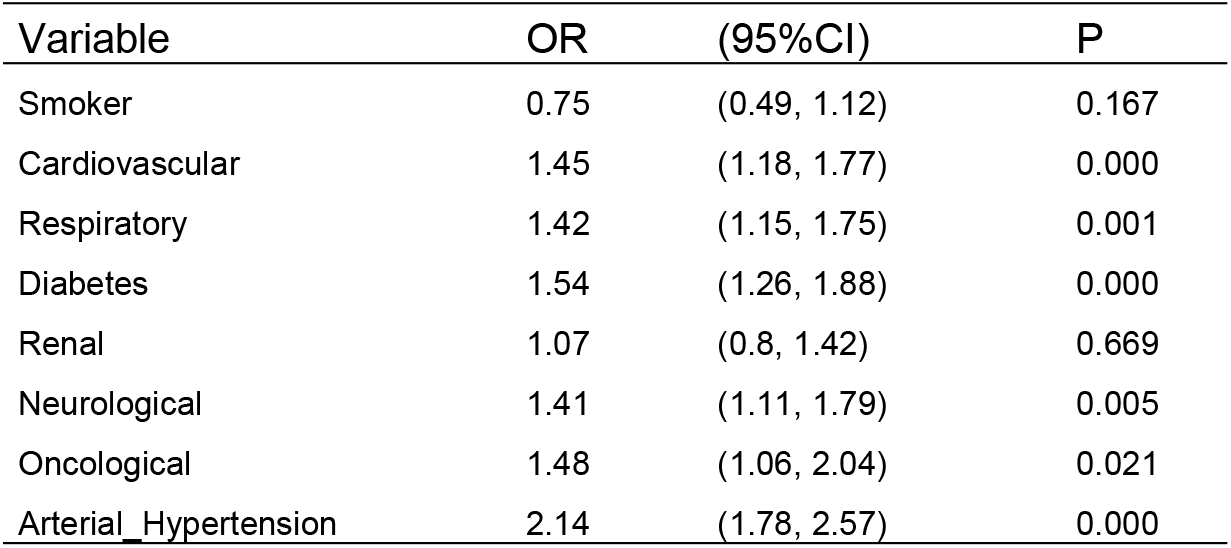
Multivariable logistic regression model for the prediction of death including all comorbidities. Note that renal morbidity and tobacco did not reach statistical significance.

| Variable | OR | (95%CI) | P |
| --- | --- | --- | --- |
| Smoker | 0.75 | (0.49, 1.12) | 0.167 |
| Cardiovascular | 1.45 | (1.18, 1.77) | 0.000 |
| Respiratory | 1.42 | (1.15, 1.75) | 0.001 |
| Diabetes | 1.54 | (1.26, 1.88) | 0.000 |
| Renal | 1.07 | (0.8, 1.42) | 0.669 |
| Neurological | 1.41 | (1.11, 1.79) | 0.005 |
| Oncological | 1.48 | (1.06, 2.04) | 0.021 |
| Arterial_Hypertension | 2.14 | (1.78, 2.57) | 0.000 |

## RESULTS

### Cohort Description

**Supplementary Table 2** shows descriptive statistics including epidemiologic data and comorbidities of the 4712 case dataset (comprising patients discharged from the ED and those requiring admission), with a global mortality rate of 13,3%. **Supplementary Table 3** describes the dataset of hospitalized patients, in whom mortality rate reached 20%. **Supplementary Figure 1** represents smoking habits and associated mortality. In spite of the fact that tobacco seemed to have a protective effect, as could be inferred from the results of the NN and PCA (**Figure 1)**, significance was lost after performing a multivariate model with all relevant variables (**Table 1**). As shown in the table, all comorbidities were found to increase mortality risk.

**Figure 1.**
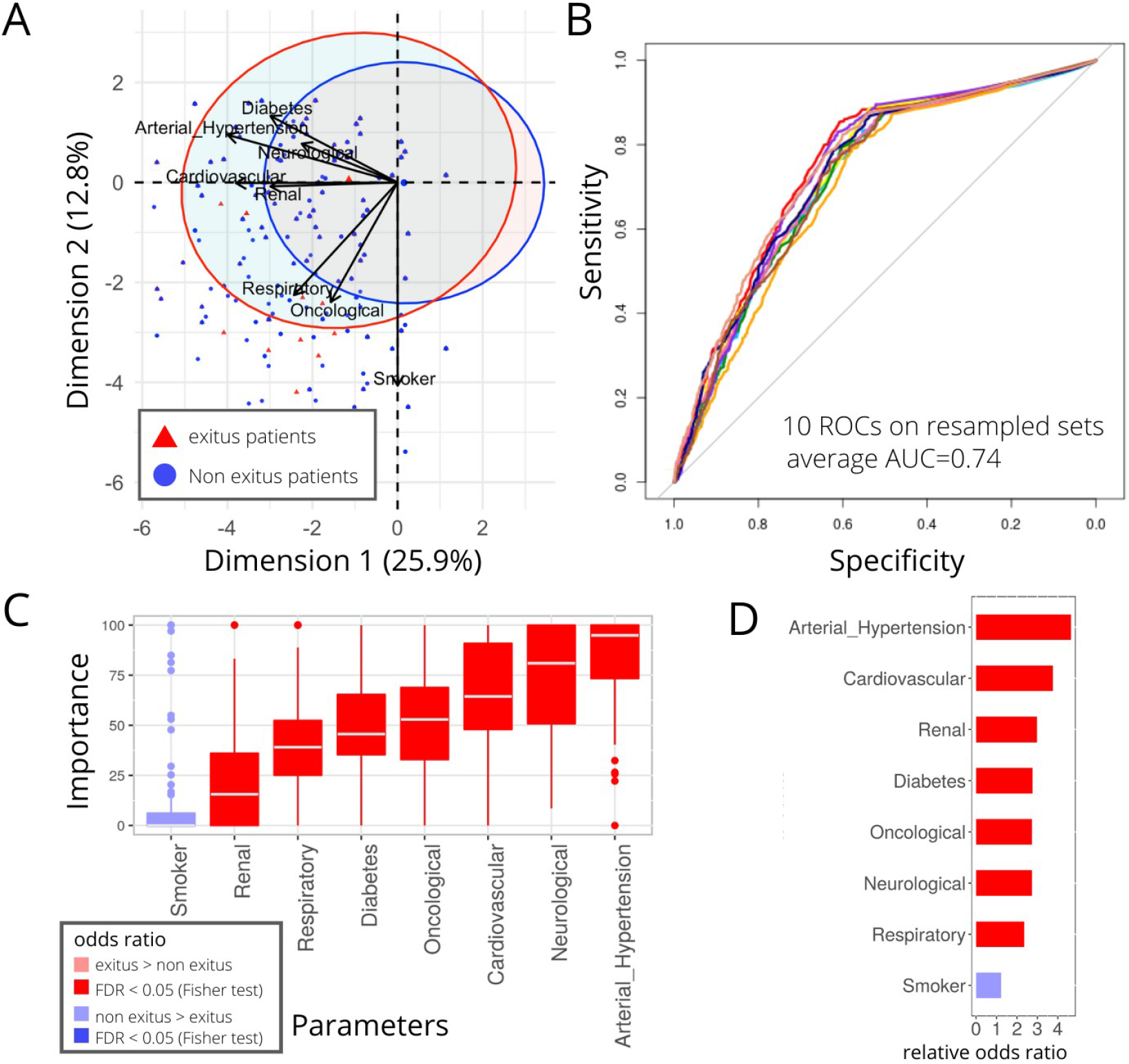
Comorbidities in Exitus and non-exitus patients of all ages. Analysis of 4712 patients. A) PCA analysis. B) Neural Network ROCs. C) Comorbidities importance in NN model. D) Fisher test results, comorbidities sorted by relative odd ratios. Significance taken as FDR<0.05.

### Comorbidities associated with worst prognosis

We performed PCA and NN analyses over the 4712 patients, classified into two states - deceased and alive-at the time of data collection. **Figure 1A** shows principal component (PC or dimension) 1 against PC2. Arterial hypertension and cardiovascular affections were the conditions which most contributed to separation between states. Of note, the NN was able to predict the status with an AUC=0.74 (accuracy=0.7, sensitivity=0.4, specificity=0.81), as can be observed in **Figure 1B**. The comorbidities which principally contributed to the NN model prediction power are shown in **Figure 1C**. As can be observed in the figure, also using this approach arterial hypertension as well as neurological and cardiovascular diseases were strongly associated to a poor outcome. Odds ratios of the different comorbidities further revealed that seven disease families were significantly enriched in patients with a fatal outcome (FDR<0.05) (**Figure 1D**). On the other hand, although smoking appeared to be more frequent between survivors, the difference was not significant.

### Risk prediction by vital signs and their cut-off values

In relationship with abnormal vital signs in the emergency department, blood pressure and temperature were found to be the variables which better discriminated between deceased patients and survivors, according to PCA analysis (**Figure 2A**). The NN was able to classify patients with an AUC=0.87 (accuracy=0.8, sensitivity=0.7, specificity=0.69, **Figure 2B**), further reaching a value of 0.91 when classification was made between deceased and ICU-admitted patients (data not shown). In addition, temperature, age and maximal heart rate were associated with death when the two latter conditions were compared (**Figure 2C)**. On the other hand, female sex, the absence of fever, adequate saturations at admission, as well as lower diastolic arterial pressures associated with survival (**Figure 2C**). In order to identify predictive cut-off values of these variables, univariate and multivariate linear regression models were carried out (**Supplementary Table 4)**. Using these thresholds, we performed a multivariate logistic regression analysis, in which age >70 years, a minimal oxygen saturation <86%, heart rate >100 bpm or fever >37.8° indicated lower survival (**Table 2)**. The AUC (CI95) for the logistic model (**Supplementary Figure 2**) was 0.87 (0.86, 0.89) and the Hosmer-Lemeshow test reached p = 0.317.

**Figure 2.**
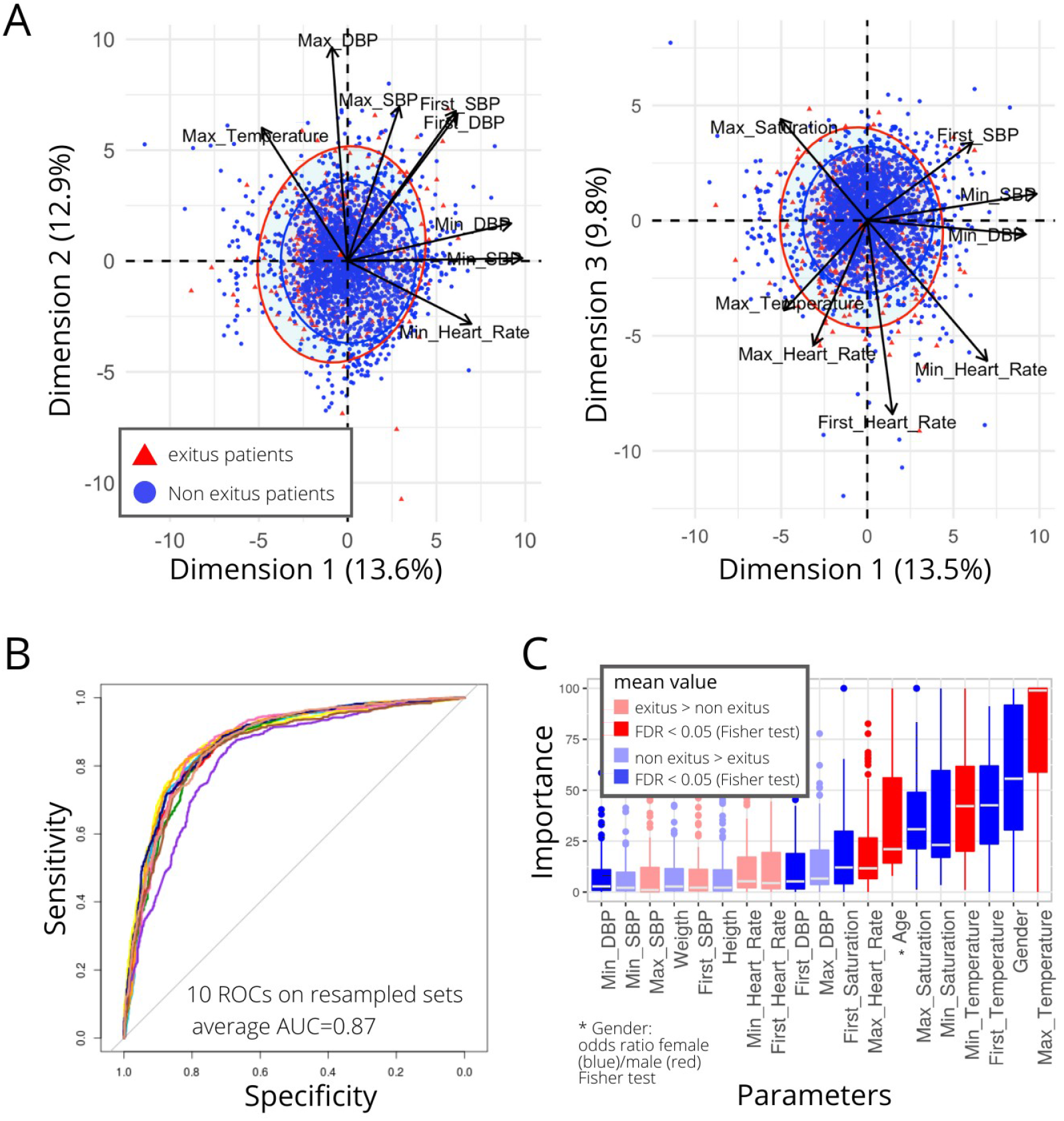
Vital signs in dead and non-dead patients of all ages. Analysis of 4712 patients. **A)** PCA analysis, Dimensions 1, 2 and 3 are shown. **B)** ROCs for Neural Network prediction. **C)** Vital signs importance in NN performance. Colors show survival or non-survival associations. Significance is calculated using a Wilcoxon test with FDR correction over p-values, FDR<0.05 was taken as significant.

**Table 2.** Multivariable model using variables grouped according to cut-off point.

| Variable | OR | (95% CI) | P |
| --- | --- | --- | --- |
| Age | 9.03 | (7.01, 11.7) | <0.001 |
| Min_Saturation | 6.43 | (4.98, 8.32) | <0.001 |
| Max_Saturation | 1.65 | (1.29, 2.12) | <0.001 |
| First_Heart_Rate | 1.29 | (1.01, 1.65) | 0.040 |
| Max_Heart_Rate | 2.56 | (1.97, 3.31) | <0.001 |
| Min_Temperature | 1.67 | (1.31, 2.13) | <0.001 |
| Max_Temperature | 1.71 | (1.33, 2.18) | <0.001 |
| Min_DBP | 0.73 | (0.56, 0.93) | 0.011 |
| Max_DBP | 2.03 | (1.58, 2.64) | <0.001 |

AUC (95%CI) = 0.87 (0.86, 0.89)
Hosmer-Lemeshow test: p = 0.317

**Table 3.**
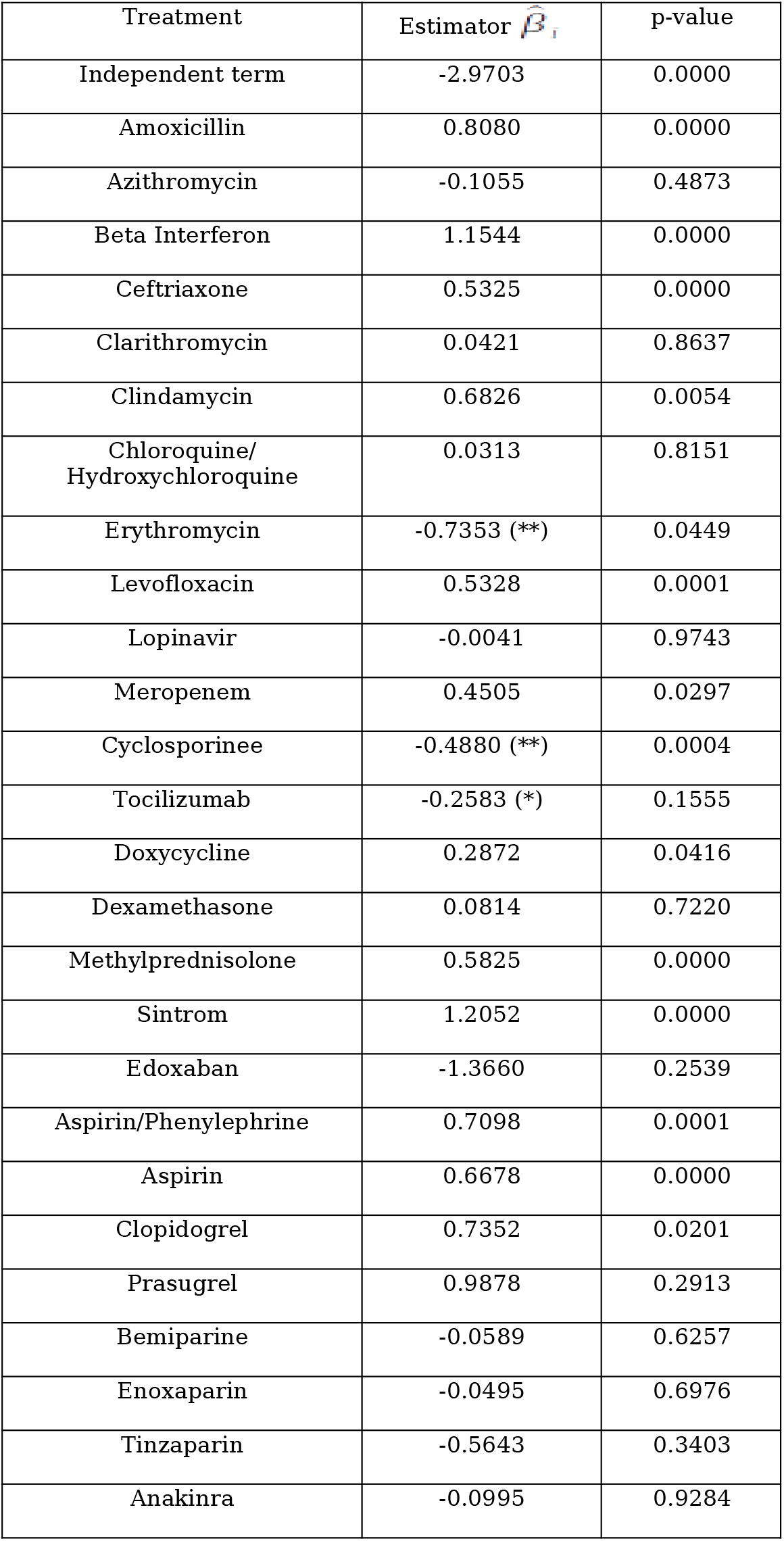
Univariate logistic regression of the relationship between each of the treatments and survival. We consider 26-predictor variables *X_i_* for each of the 26 treatments. We show the estimation *β_i_* of the coefficient *β_i_* for each of the treatments, and, in order to test statistical significance, we also show the p-value for testing *β_i_* = 0 against *βi* ≠ 0. A negative coefficient indicates that the corresponding treatment, according to the adjusted model, diminishes *p* thus being associated with survival. For the usual confidence levels, the significantly negative coefficients corresponding to a better survival were the treatments Eritromycine and Cyclosporinee, labeled (**) in the table, while Tocilizumab, labeled (*) exhibits an intermediate significance level around 15%.

| Treatment | Estimator $\hat{\beta}_i$ | p-value |
| --- | --- | --- |
| Independent term | -2.9703 | 0.0000 |
| Amoxicillin | 0.8080 | 0.0000 |
| Azithromycin | -0.1055 | 0.4873 |
| Beta Interferon | 1.1544 | 0.0000 |
| Ceftriaxone | 0.5325 | 0.0000 |
| Clarithromycin | 0.0421 | 0.8637 |
| Clindamycin | 0.6826 | 0.0054 |
| Chloroquine/<br>Hydroxychloroquine | 0.0313 | 0.8151 |
| Erythromycin | -0.7353 (**) | 0.0449 |
| Levofloxacin | 0.5328 | 0.0001 |
| Lopinavir | -0.0041 | 0.9743 |
| Meropenem | 0.4505 | 0.0297 |
| Cyclosporinee | -0.4880 (**) | 0.0004 |
| Tocilizumab | -0.2583 (*) | 0.1555 |
| Doxycycline | 0.2872 | 0.0416 |
| Dexamethasone | 0.0814 | 0.7220 |
| Methylprednisolone | 0.5825 | 0.0000 |
| Sintrom | 1.2052 | 0.0000 |
| Edoxaban | -1.3660 | 0.2539 |
| Aspirin/Phenylephrine | 0.7098 | 0.0001 |
| Aspirin | 0.6678 | 0.0000 |
| Clopidogrel | 0.7352 | 0.0201 |
| Prasugrel | 0.9878 | 0.2913 |
| Bemiparine | -0.0589 | 0.6257 |
| Enoxaparin | -0.0495 | 0.6976 |
| Tinzaparin | -0.5643 | 0.3403 |
| Anakinra | -0.0995 | 0.9284 |

### Therapeutic modalities and patient survival

#### Model 1: General logistic regression analysis: independent effects

The results are displayed in **Table 3**:

For the usual confidence levels, the significantly negative coefficients corresponding to a higher survival probability were shown by treatment with erithromycin and Cyclosporine A, labeled (**) in the table, while tocilizumab, labeled (*), exhibited an intermediate significance level around 15%. It should be mentioned, however, that erithromycin was only administered to 82 patients - mostly in intensive care as prokinetic drug-, which is a relatively small number considering the sample size.

#### Model 2: NN treatment survival prediction

In order to provide a complementary decision tool for treatment prediction of outcomes, we analysed with NN results on survival yielded by the different treatments employed in 2776 patients. The NN algorithm was applied to the age categories specified in Materials and Methods and patients were classified as dead or alive. Considering the population as a whole, the NN reached a moderate AUC=0.65 (**Figure 3A**), whereas the analysis by age categories drew a higher AUC in middle age ranges: 31-50 years, AUC=0.81 (built with 393 patients) (**Figure 3B**) and 51-70, AUC=0.67 in 1029 patients (**Figure 3C**). On the other hand, prediction capacity fell in the range of 71-100 years, with 1309 patients included and an AUC=0.59.

**Figure 3.**
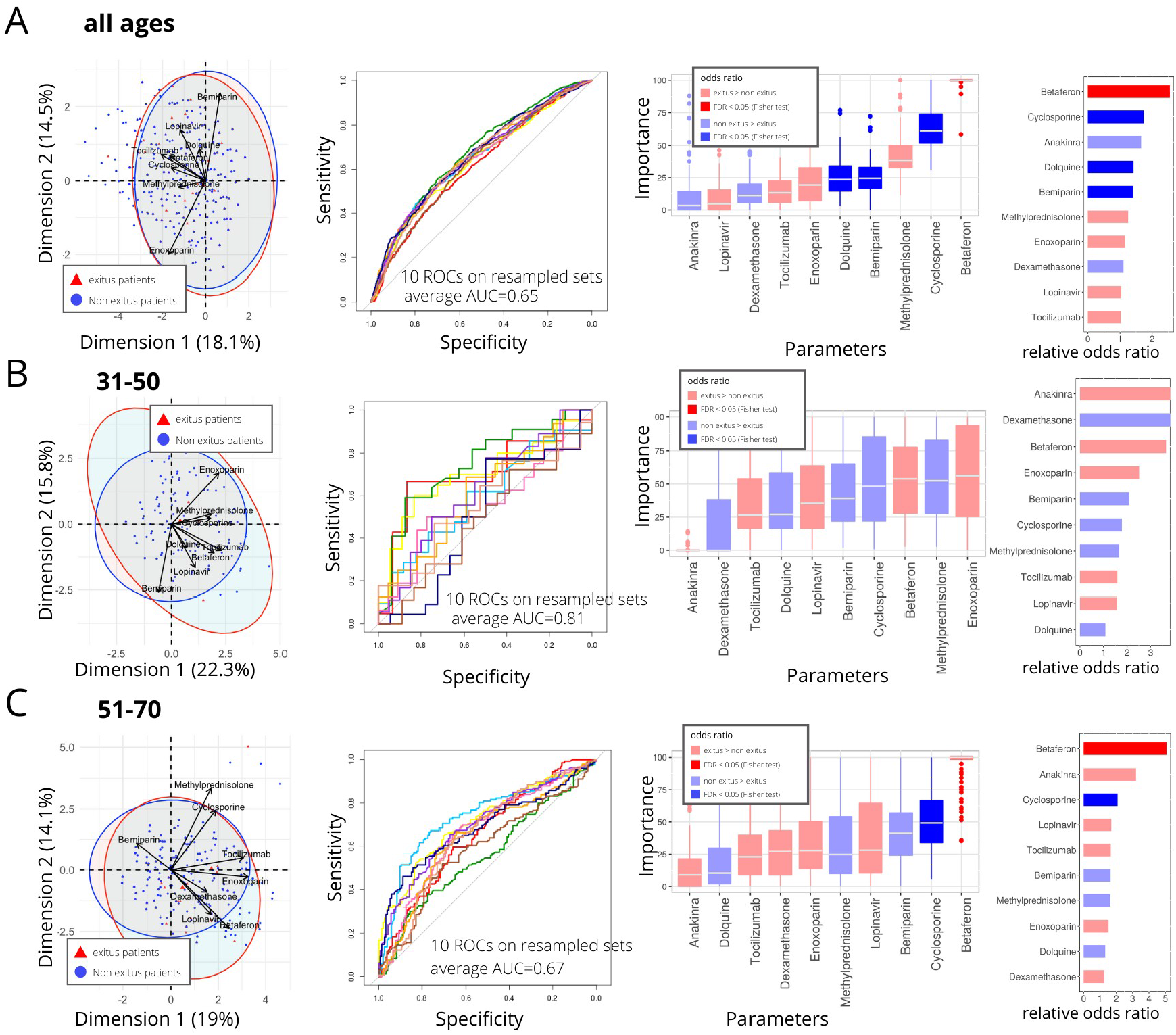
Treatments in dead and non-dead patients of three different age groups. **A)** PCA, Neural Network ROC and importance, and Fisher test results for patients of all ages (2776 patients). **B)** PCA, Neural Network ROC and importance, and Fisher test results for patients in 31-50 age range (393 patients). **C)** PCA, Neural Network ROC and importance, and Fisher test results for patients in 51-70 age range (1029 patients).

In the whole population, Cyclosporine, hydroxycloroquine, bemiparin, dexamethasone and anakinra associated to survival, albeit only the first three drugs showed a significant (FDR<0.05) enrichment in survivors (**Figure 3A**). In the range of 31-50 years old (N=393 patients) PCA showed not consistent ROC curves and no drugs associated to any of the outcomes with significant support. As regards ages between 51 and 70 years (1029 patients) **(Figure 3C)**, Cyclosporinee was the only drug associated with survival (FDR<0.05). We further explored the effect of drugs on survival in the most severe group of patients, as defined either by admission to the ICU or by a fatal outcome. Interestingly, PCA showed a clear separation between these categories and revealed a positive contribution of all drugs to survival (**Figure 4A**). NN showed an AUC=0.9 (accuracy=0.84, sensitivity=0.67, specificity=0.73, **Figure 4B**) with enoxoparin, tocilizumab and dexamethasone as the compounds with higher capacity to classify patients (**Figure 6C**). The enrichment analysis showed the benefit of those three drugs - with high odds ratios and FDR<0.05-along with an effect of lopinavir, betaferon, Cyclosporine and methylprednisolone (**Figure 4D**).

**Figure 4.**
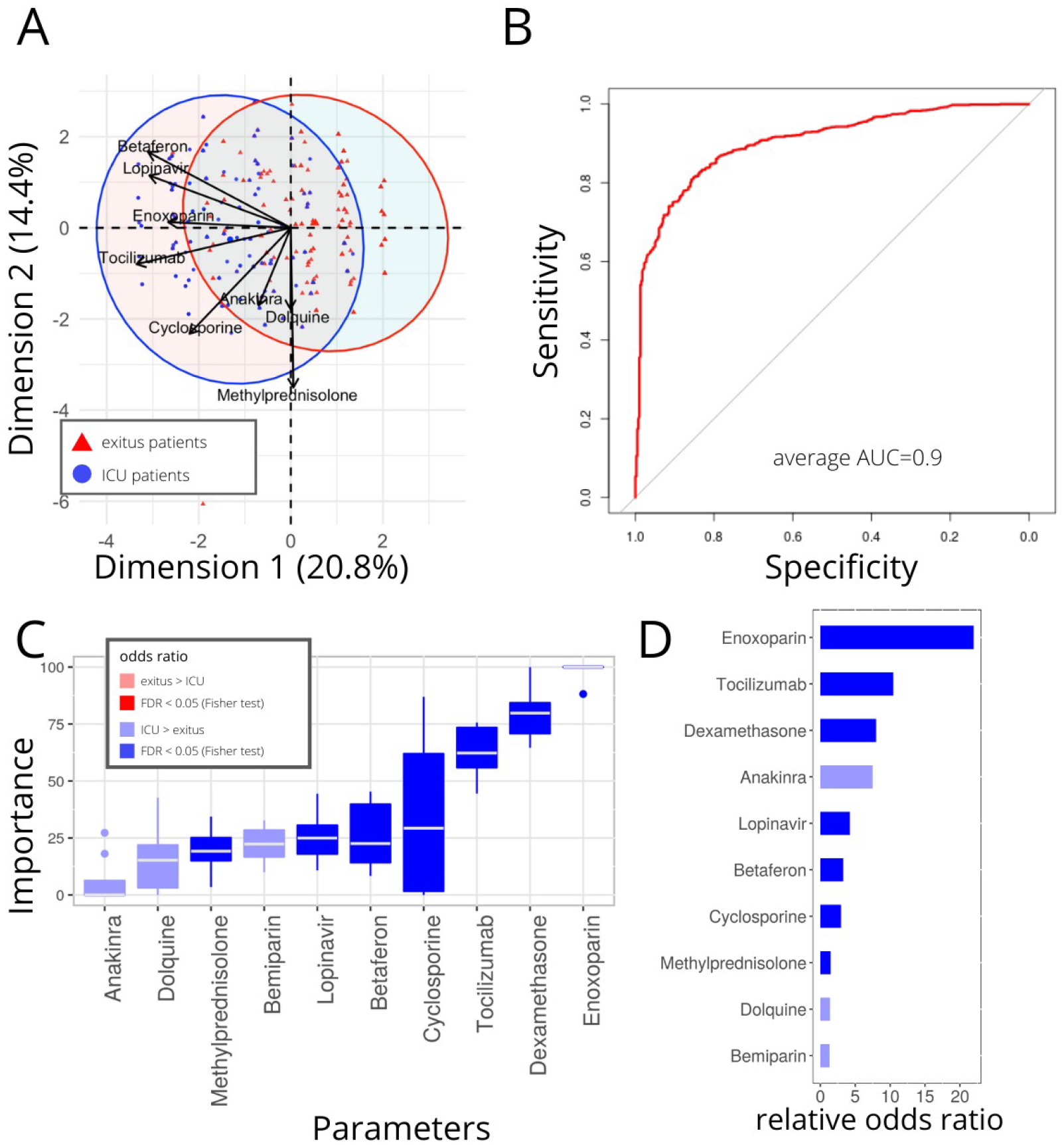
Treatments in deacesed and ICU admittedpatients of all ages. Analysis of 2776 patients. **A)** PCA analysis. **B)** NN ROCs. **C)** Drug importance in NN model. **D)** Fisher test results, drugs sorted by relative odd ratios. Significance taken as FDR<0.05.

### NN and biomarkers

Blood tests including levels of relevant analytes at first day of admission were recorded in all patients with < 50% missing data, reaching a total of 2,682 patients included in the analysis. According to PCA, three groups of biomarkers could syntethise class differences, a. monocyte and lymphocyte counts, b. urea and creatinine and c. neutrophils, segmented cell percentage and LDH (**Figure 5A**). The specific NN model yielded an AUC=0.73 (accuracy=0.69, sensitivity=0.62, specificity=0.66, **Figure 5B**). The variables with higher weights in class predictions were lymphocytes, hemoglobine and urea (**Figure 5C**). Wilcoxon rank sum test was calculated comparing the values of each variable in deceased and living patients (**Figure 5D**). While higher counts of lymphocytes, monocytes, platelets and red cells, as also levels of alanine aminitransferase and hemoglobin were found to associated with survival in this analysis, additional parameters were increased in the deceased patients. Out of them, absolute numbers of lymphocytes showed the higher discrimination capacity between outcomes.

**Figure 5.**
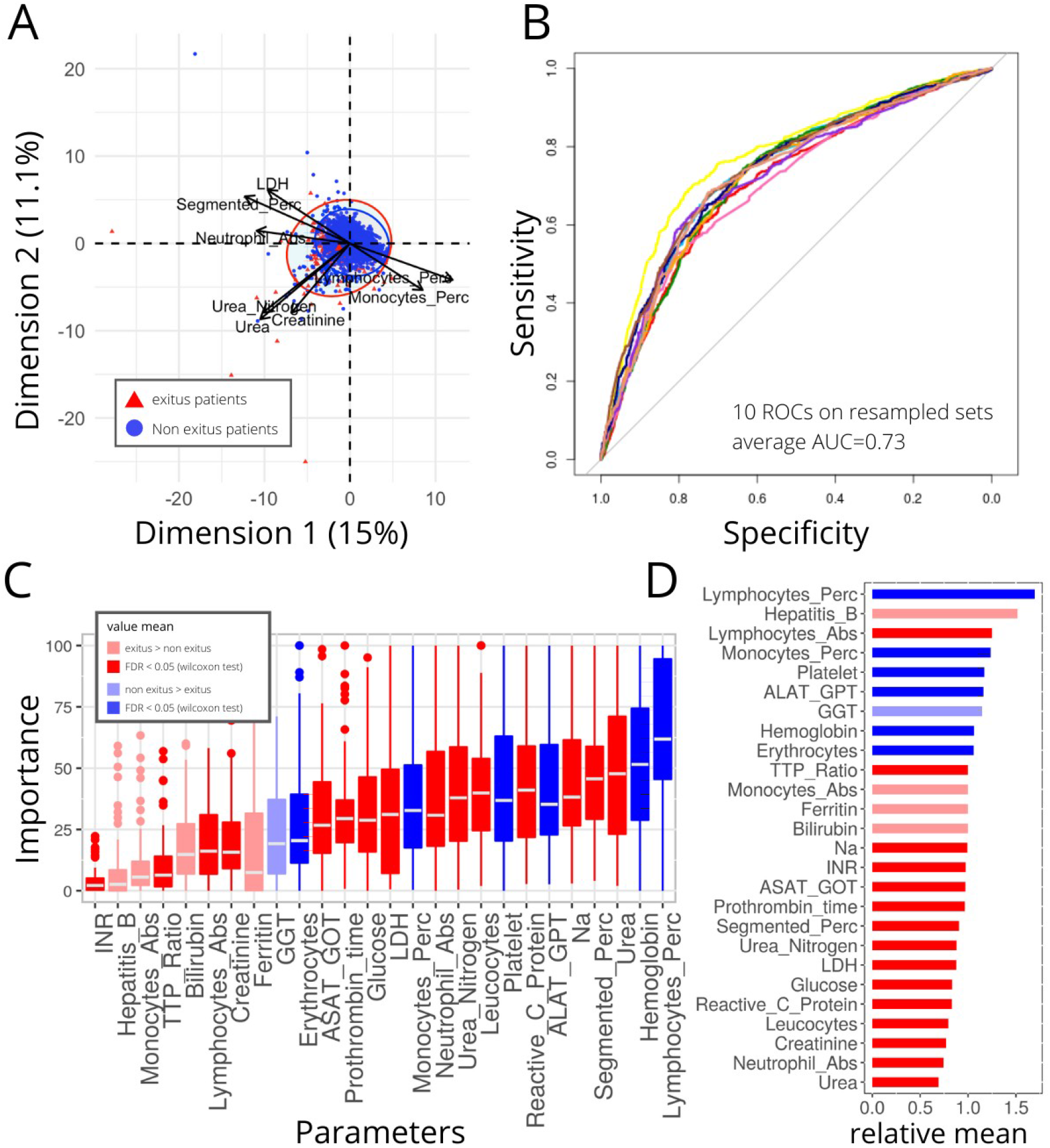
Biomarkers in dead and non-dead patients of all ages. Analysis of 2682 patients. **A)** PCA analysis. **B)** ROCs for Neural Network prediction. **C)** Biomarkers importance in NN performance. **D)** Biomarkers mean value comparison in dead and living patients, Wilcoxon test results. FDR<0.05 is taken as significant.

## DISCUSSION

The present study shows performance of a NN capable of identifying and classifying mortality predictors in SARS CoV-2 infected patients. The NN code is offered to the scientific community, since the algorithm has the ability to adapt and self-learn, accounting for a valuable predictive tool in a pandemic which is still evolving. A local clinical practice-based guidance for inpatient care was developed during the first weeks of the pandemic and subsequently subject to adaptations as specified in **Supplementary Table 1**. In accordance, criteria for admission, follow-up laboratory test profiles and therapeutic recommendations were uniform between the 4 centers, although adherence to this protocol could vary.

Regarding comorbidities, in agreement with previous data, a past history of arterial hypertension, neurological disorders and diabetes weighed negatively in the outcome of our patients. These comorbidities have been reported in other series^6,7^ which suggest that the virus preferentially targets patients with the so-called metabolic syndrome. In relation to neurological disorders, many authors have argued for a possible tropism of the virus to neural tissue, as indicated by the high prevalence of anosmia and ageusia^8^, but also by frequent cases presenting with encephalitis^9^ and strokes^10^. Intriguingly, smoking has not been found to increase risk or severity. Moreover, there are some data pointing to its possible protective role, an effect that has being claimed to rely on nicotine^11^. However, our results could not consistently support this potential effect. Targeted studies are warranted to definitively clarify this aspect, which can significantly affect public health.

Regarding clinical signs, fever>37,8°, age>70 and heart rhythm disturbances, were predictors of poor prognosis in our cohort. With respect to heart rhythm, there is certain evidence pointing to an arrhythmogenic potential of the virus^12^. Older age has repeatedly been regarded as a decisive factor in the development of severe forms of COVID19, a fact that should be taken into consideration in the distribution of resources^13^. Along with the weight of age, our study also replicated the protective effect of female gender found in other studies. In addition, we could define a range of “safe” oxygen saturations between values of 86% to 97,5%, suggesting that not only hypoxia but also hyperoxia^14^ could be detrimental.

Even though data recently disclosed from ongoing clinical trials point to the potential benefit of some compounds in the medical treatment of SARS-CoV2 pneumonia, solid evidence of efficacy of the drugs which are being used as standard therapy is still lacking^15^. At the beginning of the pandemic there was an unmet need in recommendations laid by most national administrations with respect to the use of immunomodulatory drugs. However, as had already been observed during the SARS-CoV epidemic outbreak of 2003, the development of acute respiratory failure appears to be the consequence of a vigorous innate immune response and does not exactly correlate with viral load^16^. Indeed, severe cases of SARS-CoV2 pneumonia can be distinguished for the increase in circulating levels of IL10, IL6 and TNF➔, which in association to the rise in ferritin serum concentration point to macrophage activation syndrome as the driver of acute respiratory distress and sepsis. ^17^. This insight has led to the incorporation of anti-cytokine drugs, such as tocilizumab, and Janus kinases inhibitors to clinical studies and treatment guidelines, while conversely the use of glucocorticoids remains controversial^18^.

Cyclosporine A is an immunomodulatory drug included in the WHO Essential Medicines List. The rationale for selecting this compound has been recently discussed^19^. Briefly, the presence of betacoronavirus inside immunocompetent cells is thought to induce the unfolded protein response jeopardizing peptide quality control activities and mitochondrial function^20^. Cyclosporine is able to modulate the unfolded protein response and rescue cells from irreversible damage under hypoxia and stress conditions^21^. In addition, synthetic activities of this family of virus have been shown to rely on their binding of cyclophilins, which are selectively blocked by Cyclosporine^22^, a mechanism accounting for the broad antiviral activities displayed by the drug^23^.

Remarkably, when we analysed the drugs used by our group, survival always appeared to be related to the exposure to Cyclosporinee both at the individual level and in combination. The same tendency was found using logistic regression or NN in the different age strata, both in the global cohort and more firmly in the inpatient subgroup. For all these reasons and in view of these results and some other unpublished observations we set up a randomized clinical trial assessing efficacy of Cyclosporine as add-on therapy with standard of care, which is now ongoing^24^.

As shown in **Supplementary Table 1**, the use of corticosteroids was progressively introduced in the management of our patients. Of note, patients in intensive care units were more likely to receive a prolonged use of dexamethasone. Tocilizumab was employed as rescue therapy according to the recommendations laid by the Spanish Health Ministry. Only recently have we started to use anakinra in combination to standard of care in those patients showing a poor clinical course either in case of limitation of therapeutic efferorts or in substitution of the second tocilizumab infusion.

Corticosteroids alone did not show benefit in our series, a finding that at first sight could argue against their use in COVID-19 pneumonia in accordance with current WHO guidance^25^. However, in agreement with previous observations^26^, we found that their combination with other drugs was consistently associated to a higher survival of our critically ill cases, as can be concluded from the results of the NN in Figure 4 where they stand as second therapy after tocilizumab in increasing chance of survival. Tocilizumab is currently undergoing a clinical trial, but preliminary results published as a press release^27^ and also data from a pilot study^28^ suggest that it could have a beneficial effect in this viral pneumonia.

Notably, we were able to observe that most of the specific therapies in our protocol could improve survival. In particular, given that it is the ICU admitted subgroup of patients the one exposed to a larger combination of drugs, in relationship with severity and use of maximum therapeutic effort, we explored the impact of medications in these patients. Figure 4 specifically shows patients who were admitted to ICU and survived, albeit 40 of these patients have not been discharged yet. In order of importance, the 5 drugs that were able to lower mortality in critically ill cases were enoxoparin, dexamethasone, tocilizumab, Cyclosporine and bemiparin.

In relationship with analytical markers we analysed matrix yields data, which are in agreement with previous publications. In summary, the following alterations were more likely associated to death or ICU admission in order of importance: uremia, neutrophilia, hiponatremia, raised C-reactive protein, leukocytosis, elevated levels of LDH, hypoglycemia, high levels of GOT and GGT, lymphopenia, renal failure, increased ferritin, bilirubin and lengthening of clotting times. Conversely, a normal or high percentage of lymphocytes, absence of anemia or of thrombocytopenia and and normal liver tests were protective traits.

### Limitations

This study has several limitations. Firstly, data was collected from the electronic health record database. This precluded the level of detail yielded by a manual medical record review. Secondly, ICU patients do not include those patients treated in Intermediate Respiratory Care Units, which have played an important role in the care of the severely respiratory patient. Thirdly, due to the rapid increase in ICU beds in at least one of the 4 hospitals, it was not possible to adequately assign some of the beds as ICU.

## Data Availability

The datasets analyzed during the current study are available from the corresponding author on request.

## Acknowledgements

We wish to thank all the subjects included in this study for their courage in fighting this virus, each and every one of the doctors at these four hospitals for their titanic effort and also all our statisticians, mathematicians, and the entire leadership team of the participating hospitals. The work is hosted by ISCIII project COV20/00181.

## Author Contributions

Sarah Heili-Frades MD^1#^ contributed to the conception and design of the work, did the analysis, interpretation of data and wrote the manuscrip, Pablo Minguez PHD^2#^ designed the NN algorithm, performed the analysis and wrote the manuscript, Ignacio Mahillo Fernández PHD^3^, Tomás Prieto-Rumeau PHD^4^, Antonio Herrero González^5^, Lorena de la Fuente PHD^6^ performed the statistical analysis, María Jesús Rodríguez Nieto PHD^7^, Germán Peces-Barba Romero MD PHD^1^, Mario Peces-Barba^8^, Nicolás González Mangado MD PHD^1^, María del Pilar Carballosa de Miguel MD^1^,Itziar Fernández Ormaechea MD^1^, Alba Naya prieto MD^1^, Farah Ezzine de Blas MD^1^, Luis Jiménez Hiscock MD^9^, Cesar Perez Calvo MD PHD^10^, Arnoldo Santos MD PHD^10^, Luis Enrique Muñoz Alameda MD PHD^11^ contributed with a critical revision of intellectual content, Fredeswinda Romero Bueno MD PHD^12^, Miguel Górgolas Hernández-Mora MD PHD^13^, Alfonso Cabello Úbeda MD PhD^13^, Beatriz Álvarez Álvarez MD^13^, Elizabet Petkova MD^13^, Nerea Carrasco MD^13^, Dolores Martin Rios MD^14^,, Olga Sánchez Pernaute MD PHD ^12^ developed the treatment protocol and critically revised and drafted final manuscript; All authors and the COVID-FJD-TEAM approved the final version of the manuscript.

## DECLARATIONS

### Ethics approval and consent to participate

The survey is a retrospective study with de-identified medical record data. No patient management protocols have been altered due to the study. The study was approved by our Institutional Ethics Committee.

## Appendix

The components’ full names and academics degrees of the COVID-FJD TEAM are as follows:

Andrés Giménez Velando MD, Herminia Ortiz Mayoral MD, Francisco José Laso del Hierro MD, Marwan Mohamed Choukri MD, Ainhoa Izquierdo Pérez MD, Laura Núñez García MD, Pablo López Yeste, MD Marcel José Rodríguez Guzmán MD, Abdulkader El Hachem Debek MD, José Miguel Villacampa Aubá MD PhD, Ana Isabel Tejero Redondo MD, Jose Maria Milicua Muñoz MD, Lara Colino Gomez MD, Pablo Turrion Fernandez MD, Victoria Andrea Hortiguela Martin, Anxela Vidal MD, Natividad Arias Martinez MD, Jose Luis Franqueza García MD PHD, Denis Clément emmanu Robaglia MD, Juan José Paez Vargas MD, Emilia rosas Carvajal MD, Lina Polanco MD, Ana maría Ioan MD, Angela del Pino MD, Alejandro Robles Caballero MD, Rafael Alvarez-Rementería Carbonel MD PHD, Leyre Alvarez Rubio MD PHD, Montserrat Aranzubia Ruiz MD PHD, Paz Bardon Iglesias MD PHD, Cecilia Bartolomé Bartolomé MD PHD, Soledad Bellas Cotan MD PHD, Isabel Calvete Alvarez MD PHD, Charlie Cuellar Bobadilla MD PHD, Juan Carlos De La Pinta MD PHD, Maria Del Barrio Valilla MD PHD, Maribel Garcia Vega MD PHD, Natalia Hernandez Ingelmo MD PHD, Cristina Ibañez Lorente MD PHD, M Jose Jouve Mesa MD PHD, Victorino Leal Caramazana MD PHD, Fernando Lopez Arias MD PHD, Marina Maric Govorcin MD PHD, Nuria Martinez Merino MD PHD, Florencia Manzano Lorefince MD PHD, Laura Moris Pablos MD PHD, Jaime Narvez Salazar MD PHD, Lourdes Oñate Couchet MD PHD, Mayte Relaño Covian MD PHD, Javier Rodrigo Tirado MD PHD, M Luisa Ruiz Nieto MD, Paloma Santiago Paniagua MD, Miguel Vazquez Antas MD, Fei Fei Yang MD, Raquel Garcia Ortega MD, Raquel Iglesias Guitian MD, Irene Subirana Carpi MD, Laura Manzano Lozano MD, Myriam Diez Cubillo MD, Maria Garcia Dominguez MD, Maria Montes Fernandez Micheltorena MD, Monica Gimenez Hernandez MD, Marcia Lorena Cabrera Sucre MD, Lucia Aragones Quintanero MD, Ignacio Aguado La Iglesia MD, Lucas Madrid Vazquez MD, Oscar Montero Saiz MD, Philippe Peter Granacher MD, Patricio Jose Amaro Soto MD, Jose Fernandez Cantalejo Padial MD PHD, Nieves Dominguez Garrido MD PHD, Elena Heras Sanchez MD, Laura Zamora Gómez MD, Laura Prieto Pérez MD PhD, Felipe Villar Álvarez MD PhD, Irene Carrillo Acosta MD, Aws Waleed Mohammed Al-Hayani MD, Silvia Calpena Martínez MD, Marina Castellanos González MD, Marta López de las Heras MD, Ana Cordero Guijarro, Antonio Broncano Lavado, Alicia Macías Balcayo, Marta Martín García, Alfonso Campos González MD, Alberto Encinas Vicente MD PhD, Gabriel Álvarez Curro MD, Lucía Báguena Campos MD, Eduard Teixeira MD, Raúl RubioYanguas MD, Laura Isabel García Pérez MD, Francisco J Cogolludo Pérez MD, Francisco J Guerra Blanco MD, David Pérez Pérez MD, Virginia Vasallo García MD, Araly Chacón Uribe MD, Mireya Bonet Loscertales MD PhD, María Benavides Gabernet MD, Javier Bécares Martínez, Ricardo Fernández Roblas MD PhD, Jaime Esteban MD Phd, Miguel Ángel Piris Pinilla MD PhD, José Fortes Alen MD PhD, Belén Arroyo MD, Sonsoles Barrio MD, Marcela Valverde MD, Sheila Recuero MD,, Belén Zamarro MD, Mariam Vélez MD, Clara Peiró MD, Soraya de la Fuente MD, Roberto Sierra MD, Javier López Botet MD, Antonio Herranz MD, Jorge Hernández MD, Silvia Rubio MD, Luis Nieto MD, Alicia Estrella MD, Laura Castañeda MD, Jorge Polo Sabau MD, Ana Lucía Rivero Monteagudo MD, Diego Meneses MD, Marta del Palacio Tamarit MD, Elisa Ruiz Arabi MD, Ana Venegas MD, Fernando Tornero Romero MD, Victoria Torrente MD, Pilar Barrio MD, Eduardo Alonso MD, Carolina Dassen MD, Blanca Rodríguez Alonso MD, Myriam Rodríguez Couso MD, Gabriela Rosello MD, Carmen Álvaro MD, Cici Feliz MD, María José Díez Medrano MD, Camila García MD, José Luis Larrea MD, Ana Pello MD, Beatriz González MD, Tatiana Hernández MD, Nancy Sánchez MD, Otto Olivas Vergara MD, Javier Vélez MD, Susana Fraile MD, Maite Ortega MD, Lara Cantero MD, Silvana Scaletti MD, Vanessa Pérez MD, Catalina Martín MD, Teresa Stock MD, Silvia Pérez MD, Andrés Silva MD, Alberto Andrés MD, Marta Oses MD, Miguel Morante MD, Lina Martínez MD, Juliana Botero MD, Diana Fresneda MD, Yolanda Martínez MD, Aida Franganillo MD, Amalia Gil MD, Ana Belén Jiménez MD, Adrián Arapiles MD, María Cruz Aguilera MD, Rafael Rubio MD, Alicia Sánchez MD, Begoña Sánchez MD, Rocío Cardá MD, Jersy Cárdenas MD, Lina Martínez MD, Manuel de la Calle MD, Rafael Touriño MD, José Luis Larrea MD, Miguel Morante MD, Alicia Aurea MD, Marta Monsalvo MD, Iris Martínez MD, Catalina Martín MD, Andrés Silva MD, Blanca Barroso MD, Ana Salomé Pareja MD, Ángel Rodríguez Pérez MD, Raúl Fernández Prado MD, Miguel Ángel Navas MD, Alfonso Romero MD, Ana Nieto Ribeiro MD, Beatriz Giraldez MD, Carolina Gotera Rivera MD, Teresa Gómez García MD, Erwin Javier Pinillos Robles MD, Laura de la Dueña Muñoz MD, Elena Heras Recuero MD, María de los Ángeles Zambrano Chacón MD, Fernanda Troncoso Acevedo MD, Carlos López Chang MD, Elena Cabezas Pastor MD, María José Romero Valle MD, Esther Canovas Rodríguez MD, Ángel Miracle MD, Marta González Rodriguez MD, Diana Betancor MD, Nicolás González Mangado MD, Sergio Farrais Villalba MD, Gonzalo Díaz Cano MD, José Manuel Corredera Rodríguez MD, Marina Fernández Ochoa MD, Alicia Gómez-Lopez MD, José María Romero Otero MD, Laura Ortega Martín MD, Leyre Baptista Serna MD, José Antonio Esteban Chapel MD, Andrea Castro-Villacañas Farzamnia MD, Laura Esteban-Lucía MD, Ángel Martínez Pueyo MD, Hans Paul Gaebelt MD. Pilar Llamas Sillero MD PHD, Javier Pardo Moreno MD PHD, Angel Jimenez Rodríguez MD, Raquel Barba Martin MD, Guillermo Jiménez Alvarez MD, Maria Isabel Quijano Contreras MD, Raul Mesón Gutierrez MD, Arcos Campillo Javier MD, Juan Carlos Taracido Fernandez MD, Rey Biel MD, Juan Jose Gomez Moreno MD PHD, Ana Leal Orozco MD PHD, Juan Manuel Alonso Domínguez MD, Agustin Albarracin Serra Javier MD, Martinez Peromingo Maria MD, Maria José Checa Venegas, Rebeca Armenta, Sandra Pelícano Vizuete, Ana Gloria Moreno Marcos.

### TABLES

**Supplementary Table 1.**
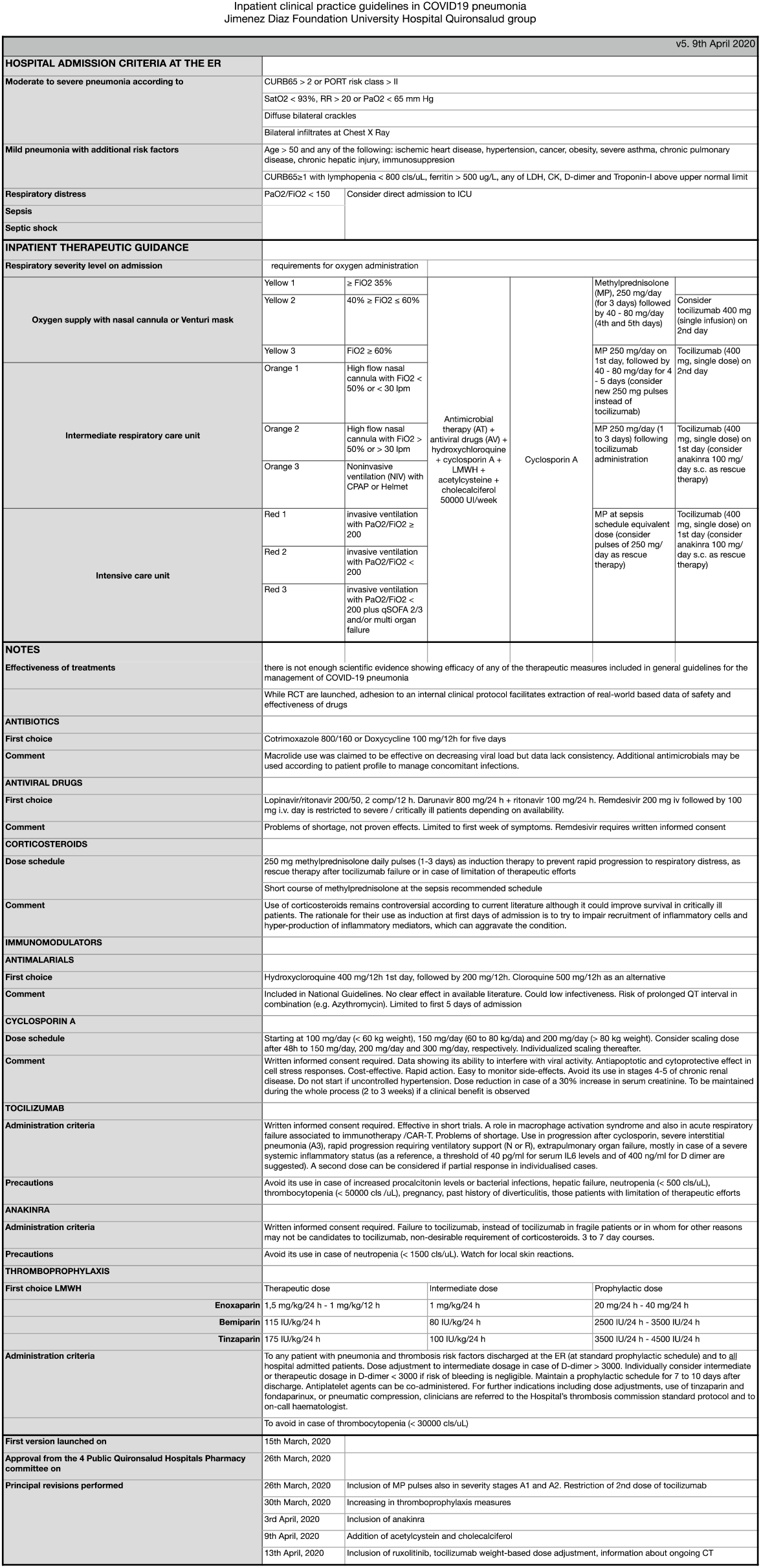
Inpatient clinical practice guidelines in COVID19 pneumonia Jimenez Diaz Foundation University Hospital Quironsalud group.

**Supplementary Table 2.** Baseline Characteristics of 4712 Patients With COVID-19 on 17 April 2020. **ICU:** intensive care unit, % of ICU: percentage of patients admitted to the ICU from the global cohort, % of Segm.: percentage referred to the segment (admitted in ICU), % of Dead: percentage of patients dead from the global cohort, % of Segm: percentage referred to the segment (Dead).

|  | Total | % | ICU | % of ICU | % of Segm. | Dead | % of Dead | % of Segm. |
| --- | --- | --- | --- | --- | --- | --- | --- | --- |
| **Demographic information** |  |  |  |  |  |  |  |  |
| Total No. | 4,712 |  | 242 |  | 5.1% | 626 |  | 13.3% |
| 0-30 | 289 | 6.1% | 3 | 1.2% | 1.0% | 1 | 0.2% | 0.3% |
| 31-50 | 1,142 | 24.2% | 41 | 16.9% | 3.6% | 14 | 2.2% | 1.2% |
| 51-70 | 1,586 | 33.7% | 159 | 65.7% | 10.0% | 97 | 15.5% | 6.1% |
| 71+ | 1,695 | 36.0% | 39 | 16.1% | 2.3% | 514 | 82.1% | 30.3% |
| Age, average | 62 |  | 60 |  |  | 81 |  |  |
| Age, standard deviation | 19 |  |  |  |  |  |  |  |
| Age, 1<sup>st</sup> quartile | 47 |  |  |  |  |  |  |  |
| Age, 3<sup>rd</sup> quartile | 77 |  |  |  |  |  |  |  |
| **Sex** |  |  |  |  |  |  |  |  |
| Female | 2,381 | 50.5% | 71 | 29.3% | 3.0% | 260 | 41.5% | 10.9% |
| Male | 2,330 | 49.4% | 171 | 70.7% | 7.3% | 365 | 58.3% | 15.7% |
| I | 1 | 0.0% | 0 | 0.0% | 0.0% | 1 | 0.2% | 100.0% |
| **Comorbidities** |  |  |  |  |  |  |  |  |
| Smoking | 1,053 | 22.3% | 70 | 28.9% | 6.6% | 184 | 29.4% | 17.5% |
| Former | 820 | 17.4% | 60 | 24.8% | 7.3% | 161 | 25.7% | 19.6% |
| Smoker | 233 | 4.9% | 10 | 4.1% | 4.3% | 23 | 3.7% | 9.9% |
| Cardiovascular | 875 | 18.6% | 33 | 13.6% | 3.8% | 256 | 40.9% | 29.3% |
| Cronic Respiratory | 687 | 14.6% | 46 | 19.0% | 6.7% | 158 | 25.2% | 23.0% |
| Diabetes | 730 | 15.5% | 53 | 21.9% | 7.3% | 180 | 28.8% | 24.7% |
| Renal | 322 | 6.8% | 12 | 5.0% | 3.7% | 97 | 15.5% | 30.1% |
| Neurological | 488 | 10.4% | 12 | 5.0% | 2.5% | 138 | 22.0% | 28.3% |
| Oncological | 239 | 5.1% | 7 | 2.9% | 2.9% | 66 | 10.5% | 27.6% |
| Hypertension | 1,794 | 38.1% | 98 | 40.5% | 5.5% | 446 | 71.2% | 24.9% |
| Metabolic disease |  |  |  |  |  |  |  |  |
| Obesity (BMI ≥30) | 607 | 12.9% | 59 | 24.4% | 9.7% | 103 | 16.5% | 17.0% |
| Morbid obesity (BMI ≥35) | 214 | 4.5% | 28 | 11.6% | 13.1% | 33 | 5.3% | 15.4% |
| **Treatment** |  |  |  |  |  |  |  |  |
| Amoxicillin | 211 | 4.5% | 14 | 5.8% | 6.6% | 72 | 11.5% | 34.1% |
| Azithromycin | 815 | 17.3% | 77 | 31.8% | 9.4% | 137 | 21.9% | 16.8% |
| Beta Interferon | 300 | 6.4% | 128 | 52.9% | 42.7% | 106 | 16.9% | 35.3% |
| Ceftriaxone | 1,563 | 33.2% | 169 | 69.8% | 10.8% | 323 | 51.6% | 20.7% |
| Clarithromycin | 131 | 2.8% | 49 | 20.2% | 37.4% | 34 | 5.4% | 26.0% |
| Clindamycin | 115 | 2.4% | 9 | 3.7% | 7.8% | 48 | 7.7% | 41.7% |
| Chloroquine / Hydroxychloroquine | 2,156 | 45.8% | 187 | 77.3% | 8.7% | 405 | 64.7% | 18.8% |
| Erythromycin | 78 | 1.7% | 76 | 31.4% | 97.4% | 21 | 3.4% | 26.9% |
| Levofloxacin | 657 | 13.9% | 98 | 40.5% | 14.9% | 172 | 27.5% | 26.2% |
| Lopinavir | 1,375 | 29.2% | 199 | 82.2% | 14.5% | 271 | 43.3% | 19.7% |
| Meropenem | 202 | 4.3% | 96 | 39.7% | 47.5% | 56 | 8.9% | 27.7% |
| Cyclosporine | 1,053 | 22.3% | 111 | 45.9% | 10.5% | 165 | 26.4% | 15.7% |
| Tocilizumab | 366 | 7.8% | 149 | 61.6% | 40.7% | 82 | 13.1% | 22.4% |
| Doxycycline | 1,177 | 25.0% | 118 | 48.8% | 10.0% | 229 | 36.6% | 19.5% |
| Dexamethasone | 194 | 4.1% | 85 | 35.1% | 43.8% | 37 | 5.9% | 19.1% |
| Methylprednisolone | 1,524 | 32.3% | 147 | 60.7% | 9.6% | 343 | 54.8% | 22.5% |

**Supplementary Table 3.**
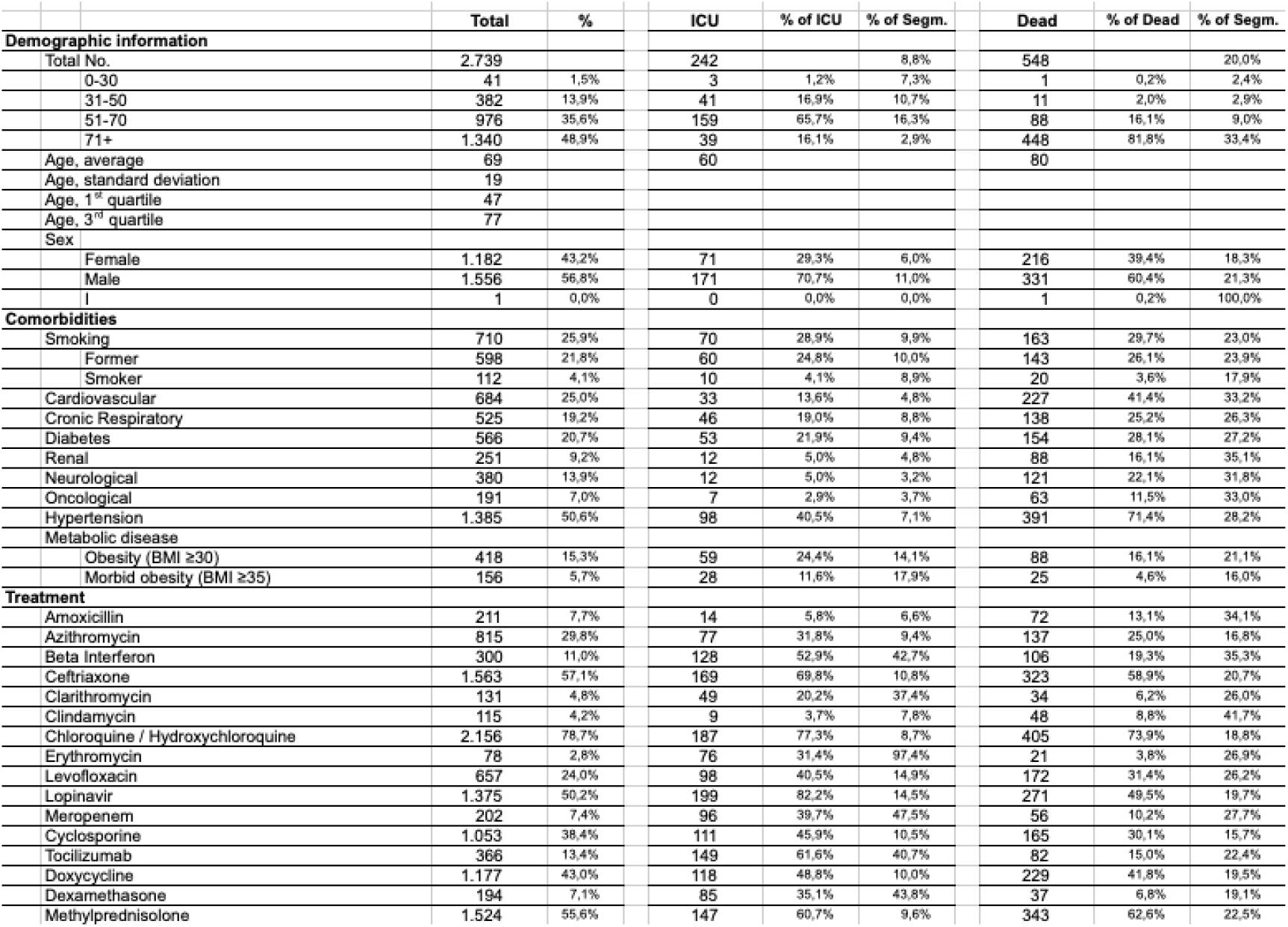
Baseline Characteristics of Patients Hospitalized With COVID-19. Baseline Characteristics of 2739 Patients Hospitalized With COVID-19 on 17th April 2020. ICU intensive care unit, % of ICU: percentage of patients admitted to the ICU from the global cohort, % of Segm: Percentage referred to the segment (admitted in ICU), % of Dead: percentage of patients dead from the global cohort, % of Segm: percentage referred to the segment (Dead)

**Supplementary Table 4.** Cutpoints for death risk determinants based on Youden’s criteria and obtained by bootstrapping.

| Variable | Cutoff | Direction | Sensibility | Specificity | AUC | P-value |
| --- | --- | --- | --- | --- | --- | --- |
| Age | 70.0 | > | 0.836 | 0.700 | 0.830 | <0.001 |
| Weigth | 77.4 | < | 0.779 | 0.245 | 0.510 | 0.559 |
| Heigth | 169 | < | 0.805 | 0.183 | 0.474 | 0.762 |
| First_Saturation | 95.0 | < | 0.804 | 0.226 | 0.410 | 0.489 |
| Min_Saturation | 86.0 | < | 0.553 | 0.900 | 0.715 | <0.001 |
| Max_Saturation | 97.5 | < | 0.650 | 0.405 | 0.439 | <0.001 |
| First_SBP | 123 | > | 0.754 | 0.357 | 0.500 | <0.001 |
| X | 105 | < | 0.746 | 0.383 | 0.480 | <0.001 |
| Max_SBP | 136 | > | 0.765 | 0.327 | 0.484 | <0.001 |
| First_DBP | 72.3 | < | 0.736 | 0.317 | 0.428 | <0.001 |
| Min_DBP | 60.0 | < | 0.474 | 0.728 | 0.573 | <0.001 |
| Max_DBP | 86.0 | < | 0.741 | 0.404 | 0.504 | <0.001 |
| First_Heart_Rate | 83.0 | > | 0.747 | 0.397 | 0.504 | <0.001 |
| Min_Heart_Rate | 64.6 | > | 0.704 | 0.442 | 0.501 | <0.001 |
| Max_Heart_Rate | 100 | > | 0.541 | 0.803 | 0.652 | <0.001 |
| First_Temperature | 37.0 | < | 0.775 | 0.310 | 0.446 | <0.001 |
| Min_Temperature | 35.7 | > | 0.736 | 0.310 | 0.453 | <0.001 |
| Max_Temperature | 37.8 | > | 0.521 | 0.763 | 0.610 | <0.001 |

### FIGURE LEGENDS

**Supplementary Figure 1.**
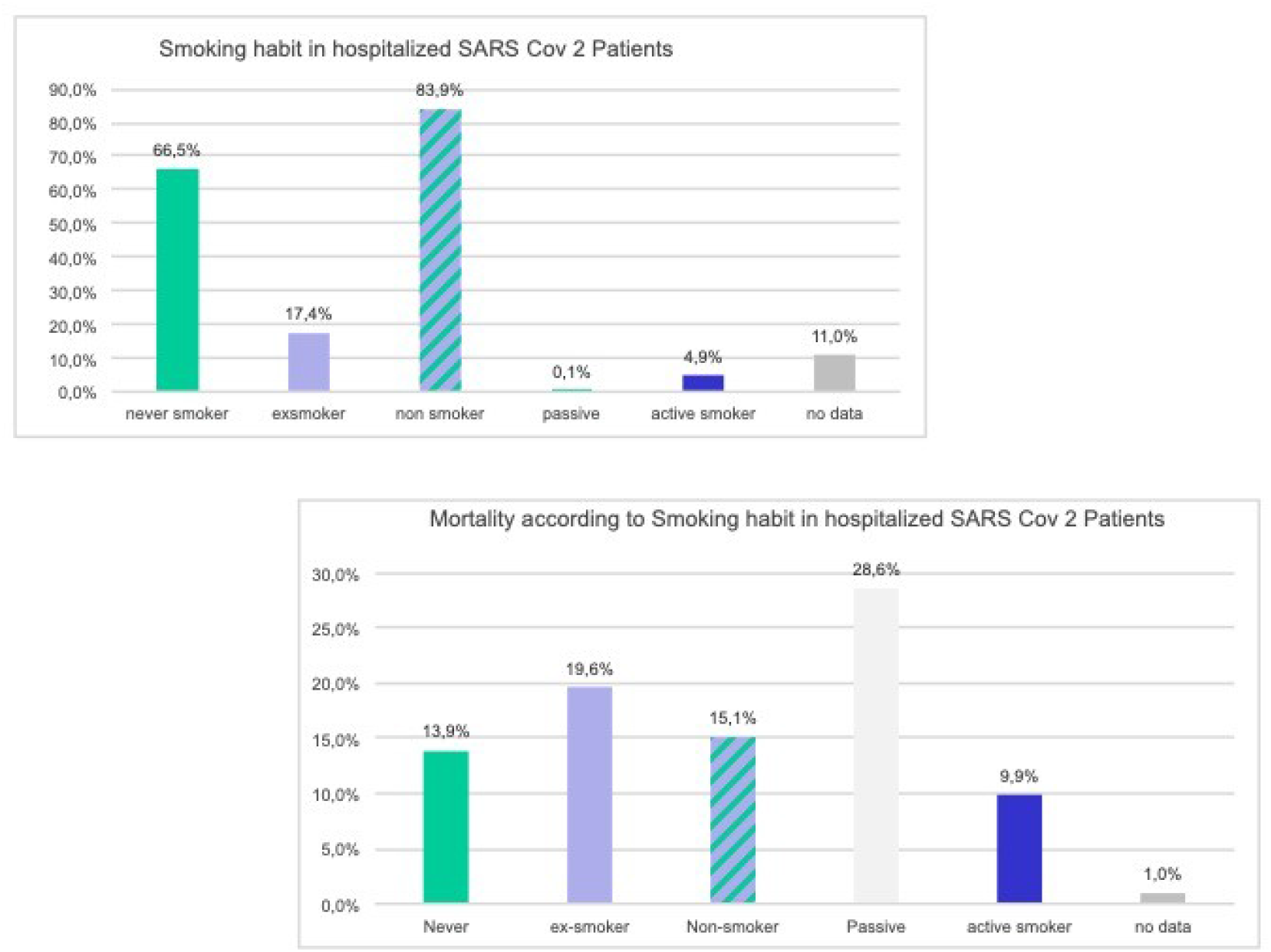
Smoking in the analyzed cohort of 4712 patients, results in terms of hospitalization and mortality.

**Supplementary Figure 2.**
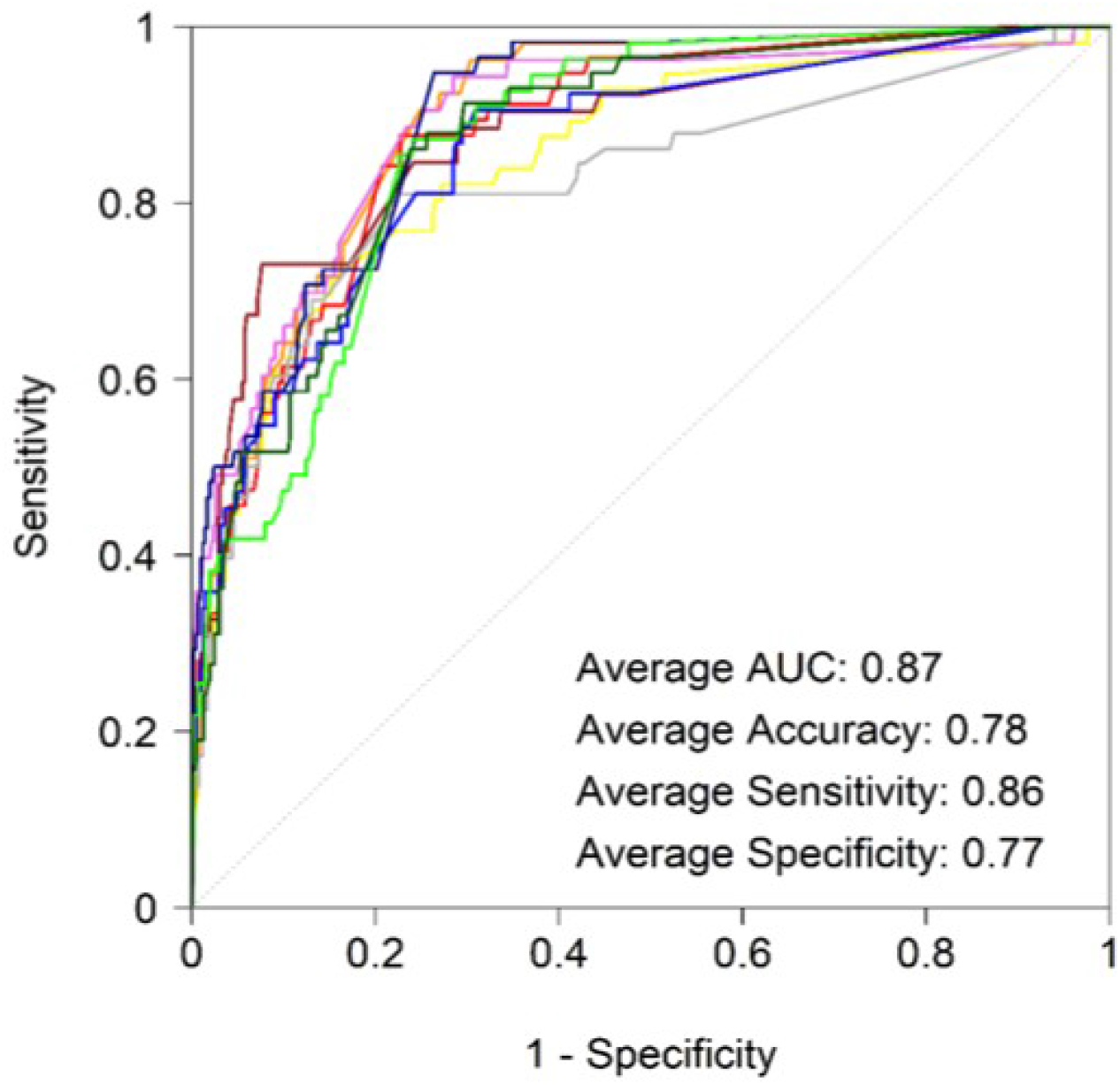
Graph with ROC curves obtained by cross validation (10 cross fold validation) for vital signs description.

## BIBLIOGRAPHY

1 Francisco Díez-Fuertes, María Iglesias-Caballero, Sara Monzón, Pilar Jiménez, Sarai Varona, Isabel Cuesta et al. Phylodynamics of SARS-CoV-2 transmission in Spain. bioRxiv 2020.04.20.050039; doi: https://doi.org/10.1101/2020.04.20.050039

2 Zhou F, Yu T, Du R, Fan G, Liu Y, Liu Z et al. Clinical course and risk factors for mortality of adult inpatients with COVID-19 in Wuhan, China: a retrospective cohort study. Lancet. 2020 Mar 28; 395(10229):1054–1062. Doi: 10.1016/S0140-6736(20)30566-3.

3 Young BE, Ong SWX, Kalimuddin S, Low JG, Tan SY, Loh J. Epidemiologic Features and Clinical Course of Patients Infected With SARS-CoV-2 in Singapore. JAMA. 2020 Mar 3. doi: 10.1001/jama.2020.3204.

4 Spina S, Marrazzo F, Migliari M, Stucchi R, Sforza A, Fumagalli R. The response of Milan’s Emergency Medical System to the COVID-19 outbreak in Italy. Lancet. 2020 Mar 14;395(10227):e49-e50. doi: 10.1016/S0140-6736(20)30493-1.

5 Richardson S, Hirsch JS, Narasimhan M, Crawford JM, McGinn. T, Davidson KW. Presenting Characteristics, Comorbidities, and Outcomes Among 5700 Patients Hospitalized With COVID-19 in the New York City Area. JAMA. 2020 Apr 22. doi: 10.1001/jama.2020.6775.

6 Huang I, Lim MA, Pranata R. Diabetes mellitus is associated with increased mortality and severity of disease in COVID-19 pneumonia - A systematic review, meta-analysis, and meta-regression. Diabetes Metab Syndr. 2020 Apr 17;14(4):395–403. doi: 10.1016/j.dsx.2020.04.018.

7 Li J, Wang X, Chen J, Zhang H, Deng A. Association of Renin-Angiotensin System Inhibitors With Severity or Risk of Death in Patients With Hypertension Hospitalized for Coronavirus Disease 2019 (COVID-19) Infection in Wuhan, China. JAMA Cardiol. 2020 Apr 23. doi: 10.1001/jamacardio.2020.1624.

8 Vaira LA, Salzano G, Deiana G, De Riu G. Anosmia and Ageusia: Common Findings in COVID-19 Patients. Laryngoscope. 2020 Apr 1. doi: 10.1002/lary.28692.

9 Ye M, Ren Y, Lv T. Encephalitis as a clinical manifestation of COVID-19.Brain Behav Immun. 2020 Apr 10. pii: S0889–1591(20)30465-7. doi: 10.1016/j.bbi.2020.04.017.

10 Carod-Artal FJ. Neurological complications of coronavirus and COVID-19. Rev Neurol. 2020 May 1;70(9):311–322. doi: 10.33588/rn.7009.2020179.

11 Brake SJ, Barnsley K, Lu W, McAlinden KD, Eapen MS, Sohal SS. Smoking Upregulates Angiotensin-Converting Enzyme-2 Receptor: A Potential Adhesion Site for Novel Coronavirus SARS-CoV-2 (Covid-19). J Clin Med. 2020 Mar 20;9(3). pii: E841. doi: 10.3390/jcm9030841.

12 Wu CI, Postema PG, Arbelo E, Behr ER, Bezzina CR, Napolitano et al. SARS-CoV-2, COVID-19 and inherited arrhythmia syndromes. Heart Rhythm. 2020 Mar 31. pii: S1547–5271(20)30285-X. doi: 10.1016/j.hrthm.2020.03.024.

13 Archard D, Caplan A. Is it wrong to prioritise younger patients with covid-19? BMJ. 2020 Apr 22; 369:m1509. doi: 10.1136/bmj.m1509.

14 S.B. Heili-Frades, E. L’Her, F. Lellouche Oxigenoterapia. Nuevos datos de toxicidad, nuevas recomendaciones y soluciones innovadoras: sistemas automatizados de titulación y destete de oxigenoterapia Rev Patol Respir. 2020; 23(1): 15–23

15 Sanders JM, Monogue ML, Jodlowski TZ, Cutrell JB. Pharmacologic Treatments for Coronavirus Disease 2019 (COVID-19): A Review. JAMA. 2020 Apr 13. doi: 10.1001/jama.2020.6019.

16 Peiris JS, Chu CM, Cheng VC, Chan KS, Hung IF, Poon LL, et al; HKU/UCH SARS Study Group. Clinical progression and viral load in a community outbreak of coronavirus-associated SARS pneumonia: a prospective study. Lancet. 2003 May 24;361(9371):1767–72.

17 Giamarellos-Bourboulis EJ, Netea MG, Rovina N, Akinosoglou K, Antoniadou A, Antonakos N, et al. Complex Immune Dysregulation in COVID-19 Patients with Severe Respiratory Failure. Cell Host Microbe. 2020 Apr 17. pii: S1931–3128(20)30236-5. doi: 10.1016/j.chom.2020.04.009.

18 Russell CD, Millar JE, Baillie JK. Clinical evidence does not support corticosteroid treatment for 2019-nCoV lung injury. Lancet. 2020 Feb 15;395(10223):473–475. doi: 10.1016/S0140-6736(20)30317-2.

19 Sanchez-Pernaute O, Romero-Bueno FI, Selva-O’Callaghan A. Why choose Cyclosporine A as first-line therapy in COVID-19 pneumonia. Reumatol Clin 2020 (in press). DOI: 10.1016/j.reuma.2020.03.001

20 Chan CP, Siu KL, Chin KT, Yuen KY, Zheng B, Jin DY. Modulation of the unfolded protein response by the severe acute respiratory syndrome coronavirus spike protein. J Virol. 2006 Sep;80(18):9279–87.

21 Du S, Hiramatsu N, Hayakawa K, Kasai A, Okamura M, Huang T, Yao J, Takeda M, Araki I, Sawada N, Paton AW, Paton JC, Kitamura M. Suppression of NF-kappaB by Cyclosporine a and tacrolimus (FK506) via induction of the C/EBP family: implication for unfolded protein response. J Immunol. 2009 Jun 1;182(11):7201–11. doi: 10.4049/jimmunol.0801772.

22 Pfefferle S, Schöpf J, Kögl M, Friedel CC, Müller MA, Carbajo-Lozoya J, Stellberger T, von Dall’Armi E, Herzog P, Kallies S, Niemeyer D, Ditt V, Kuri T, Züst R, Pumpor K, Hilgenfeld R, Schwarz F, Zimmer R, Steffen I, Weber F, Thiel V, Herrler G, Thiel HJ, Schwegmann-Wessels C, Pöhlmann S, Haas J, Drosten C, von Brunn A. The SARS-coronavirus-host interactome: identification of cyclophilins as target for pan-coronavirus inhibitors. PLoS Pathog. 2011 Oct;7(10):e1002331. doi: 10.1371/journal.ppat.1002331.

23 De Wilde AH, Raj VS, Oudshoorn D, Bestebroer TM, van Nieuwkoop S, Limpens RWAL, Posthuma CC, van der Meer Y, Bárcena M, Haagmans BL, Snijder EJ, van den Hoogen BG. MERS-coronavirus replication induces severe in vitro cytopathology and is strongly inhibited by Cyclosporine A or interferon-α treatment. J Gen Virol. 2013 Aug;94(Pt 8):1749–1760. doi: 10.1099/vir.0.052910-0.

24 The SARS-CoV2 study group of HU Fundación Jimenez Díaz is currently conducting a clinical trial to test efficacy and safety of CsA as add-on therapy with standard of care in a 1:1 randomization with severity stratification (EudraCT 2020–002621-11; ClinicalTrialsGov)

25 WHO Clinical management of severe acute respiratory infection when novel coronavirus (nCoV) infection is suspected. Jan 28, 2020. https://www.who.int/publications-detail/clinical-management-of-severe-acute-respiratory-infection-when-novelcoronavirus-(ncov)-infection-is-suspected

26 Zhou W, Liu Y, Tian D, Wang C, Wang S, Cheng J, et al. Potential benefits of precise corticosteroids therapy for severe 2019-nCoV pneumonia. Signal Transduct Target Ther. 2020 Feb 21;5(1):18. doi: 10.1038/s41392-020-0127-9.

27 https://www.aphp.fr/contenu/le-tocilizumab-ameliore-significativement-le-pronostic-des-patients-avec-pneumonie-covid

28 Zhang C1, Wu Z1, Li JW1, Zhao H2, Wang GQ3. The cytokine release syndrome (CRS) of severe COVID-19 and Interleukin-6 receptor (IL-6R) antagonist Tocilizumab may be the key to reduce the mortality. Int J Antimicrob Agents. 2020 Mar 29:105954. doi: 10.1016/j.ijantimicag.2020.105954.

